# A systematic review of anaesthetic agents used in Drug Induced Sedation Endoscopy (DISE) and a description of a new DISE technique

**DOI:** 10.1101/2021.12.02.21267209

**Authors:** Oliver Sanders, Bhik Kotecha, Vik Veer

**Affiliations:** Southend University Hospital; Royal National Throat, Nose and Ear Hospital

## Abstract

**Objectives:** Drug induced sleep endoscopy (DISE) is a standardly used investigation for surgical planning in obstructive sleep apnoea management once conservative treatments have proven inadequate. There are a variety of anaesthetic agents used to obtain sedation necessary for DISE. These agents may have different effect on the upper airway and other parameters important in the diagnosis of the site of collapse during sleep. We aimed to review the commonly agents and evaluate the significance of their impact on the the diagnosis.

**Methods:** A search was conducted through PubMed looking for studies on commonly used anaesthetic agents and their effect on the upper airway and cardiopulmonary parameters. Results: Of the 109 studies yielded by the search, 19 were deemed relevant to the review and met all inclusion criteria. The agents reviewed were: propofol, dexmedetomidine, remifentanil, isoflurane, sevoflurane, midazolam and topical lidocaine. A meta-analysis was not conducted due to the limited number of relevant studies and the heterogeneity of outcomes measured. All agents examined gave some element of airway collapse and impact on cardiopulmonary measures. Most of these effects were shown to be dose-dependent. Of the agents considered dexmedetomidine and propofol gave the most consistently reliable and physiologically safe representations of upper airway collapse seen in OSA patients.

**Conclusion:** There is limited information and no industry standard for the sedative regimen used for DISE. Of the agents reviewed those that caused least cardiopulmonary instability, respiratory depression and exaggerated upper airway collapse were deemed the most appropriate for DISE. The agent that best meet these criteria is dexmedetomidine followed by propofol.

## Introduction

Obstructive sleep apnoea (OSA) is a disease process characterised by collapse of the upper airway structures during sleep(1). Some of the pathological sequelae of OSA include hypertension, ischaemic heart disease, diabetes and depression(2).

Drug Induced Sedation Endoscopy (DISE) was first described in 1991 by Croft & Pringle (3) in our institution. A state similar to natural sleep is induced using anaesthetic agents. Flexible nasendoscope is used to examine the upper aero-digestive tract during this light sedative state to ascertain the probable location of collapse in these OSA patients(4). Therefore, the main aim of DISE is in identifying the location, or more commonly locations (5) of upper airway collapse so that targeted therapy options may be employed (6). The obvious criticism of this technique is that this is not natural sleep (7). How can one be certain that the airway collapse seen under the influence of anaesthetic agents is the same as that would have been seen during natural sleep?(8,9) Choice of sedative agent therefore is paramount, and this article systematically reviews advantages and disadvantages of the drugs used to induce a sleep-like state. The authors also introduce and describe a new method of performing a DISE that we believe responds to some of the criticism of DISE.

### Methods

The inclusion criteria this review were original data human studies, published in English, which involved more than two subjects. Inclusion also required evaluation of the upper airway or measurement of physiological parameters pertaining to upper airway dynamics. Excluded studies included any animal data and other reviews. Review articles and the references of each article gained were examined to obtain further studies not acquired during the initial literature searches.

PubMed was used with the agreed search terms

((“anesthesia”[MeSH] OR “sedation”[All Fields]) AND (“upper airway”[All Fields] OR “drug induced sedation endoscopy”[All Fields] OR “DISE”[All Fields])) AND (“humans”[MeSH Terms] AND “english”[lang]) AND (“lidocaine”[All Fields] OR “isoflurane”[All Fields] OR “sevoflurane”[All Fields] OR “desflurane”[All Fields] OR “propofol”[All Fields] OR “dexmedetomidine”[All Fields] OR “opioids”[All Fields] OR “fentanyl”[All Fields] OR “remifentanil”[All Fields] OR “alfentanil”[All Fields] OR “morphine”[All Fields] OR “midazolam”[All Fields])

This search yielded 109 studies which following screening of abstracts was reduced to 30. Full text articles were sourced and assessed. A further 11 were excluded due to unrelated data not apparent from the abstracts.

From the final 19 papers the following information was extracted: first author, year of publication, study design, number of participants, inclusion criteria, intervention, outcomes measured and conclusion of the study. Outcomes measured in the papers reviewed included:

- Polysomnography findings including AHI
- Upper airway cross sectional areas in both static and dynamic MR imaging
- Airflow dynamics on CPAP measuring Pcrit (pressure required to overcome airway obstruction)
- Electromyography of the genioglossus muscle (EMGgg)
- Observed locations of obstruction using DISE – most commonly in VOTE (velopharynx, oropharynx, tongue base and epiglottis)(10) anatomical locations
- Cardiopulmonary parameters.

It was not possible to perform a meta-analysis owing to the heterogeneity of the measured outcomes from the studies.

## Results

**Table 1.** illustrates the studies obtained for each type of anaesthetic agent.

| Anaesthetic agent | Studies reviewed |
| --- | --- |
| Propofol | • Berry et al. 2005 (Bolus)<br>• Hillman et al. 2009 (TCI)<br>• Rabelo et al. 2010 (TCI) |
|  | • Evans et al. 2003 (TCI)<br>• De Vito et al. 2011 (Bolus vs TCI) |
| Dexmedetomidine | • Mahmoud et al. 2013 (TCI)<br>• Mahmoud et al. 2009 (DEX vs Propofol)<br>• Yoon et al. 2016 (TCI)<br>• Capasso et al. 2016 (DEX Vs Propofol)<br>• Mahmoud et al. 2010 (TCI) |
| Remifentanyl | • Cho et al. 2015 (infusion) |
| Isoflurane | • Eastwood et al. 2002 (end-tidal) |
| Sevoflurane | • Crawford et al. 2006 (MAC) |
| Midazolam | • Genta et al. 2011 (bolus)<br>• Abdullah et al. 2013 (bolus) |
| Lidocaine | • McNicholas et al. 1987 (oropharyngeal vs nasal)<br>• Berry et al. 1995<br>• Ho et al. 2006<br>• DeWeese et al. 1988 |

Table 2 summarises the findings of the 19 studies reviewed and found to be appropriate grouped into the agent about which the studies was based.

**Table 2.**
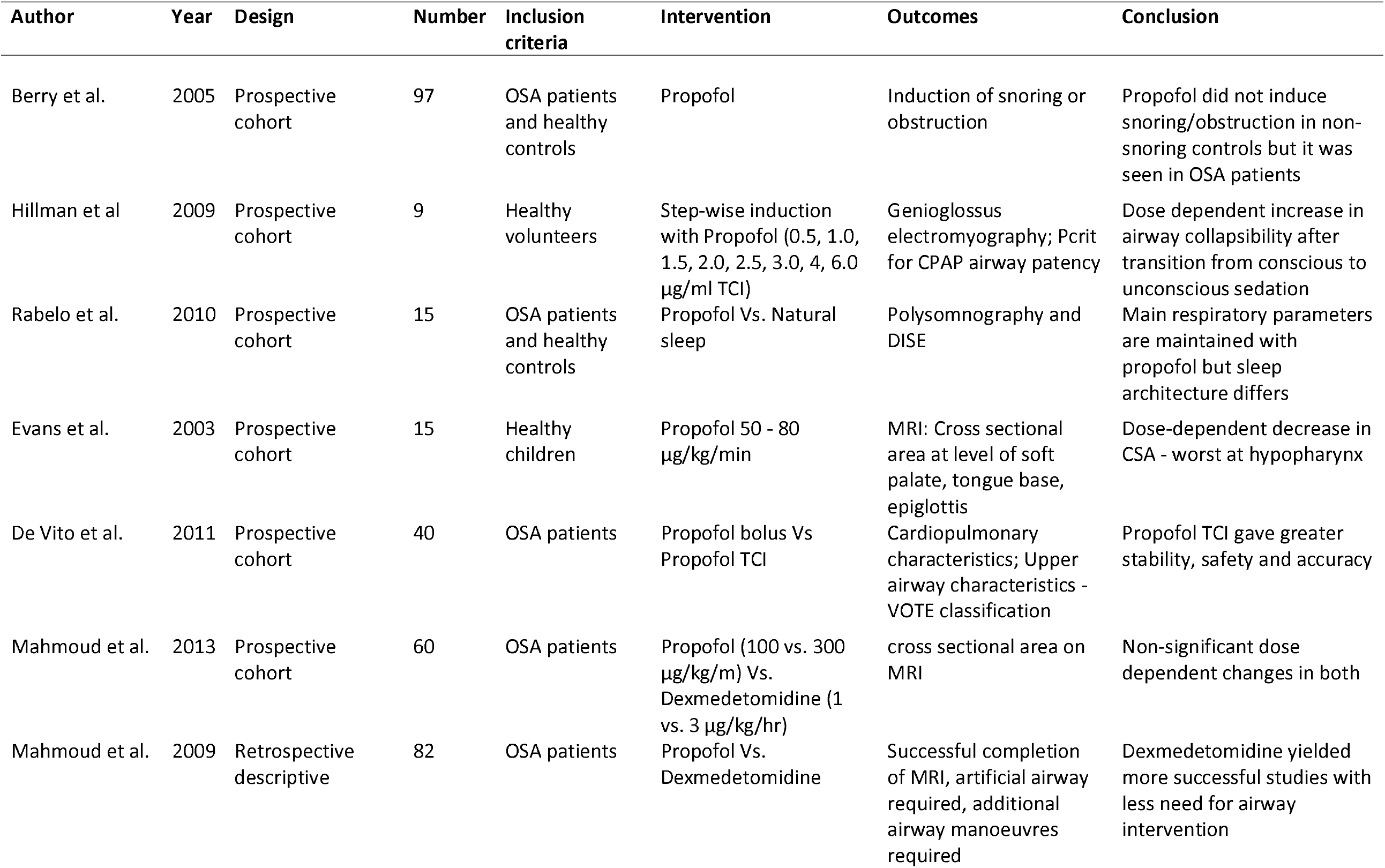
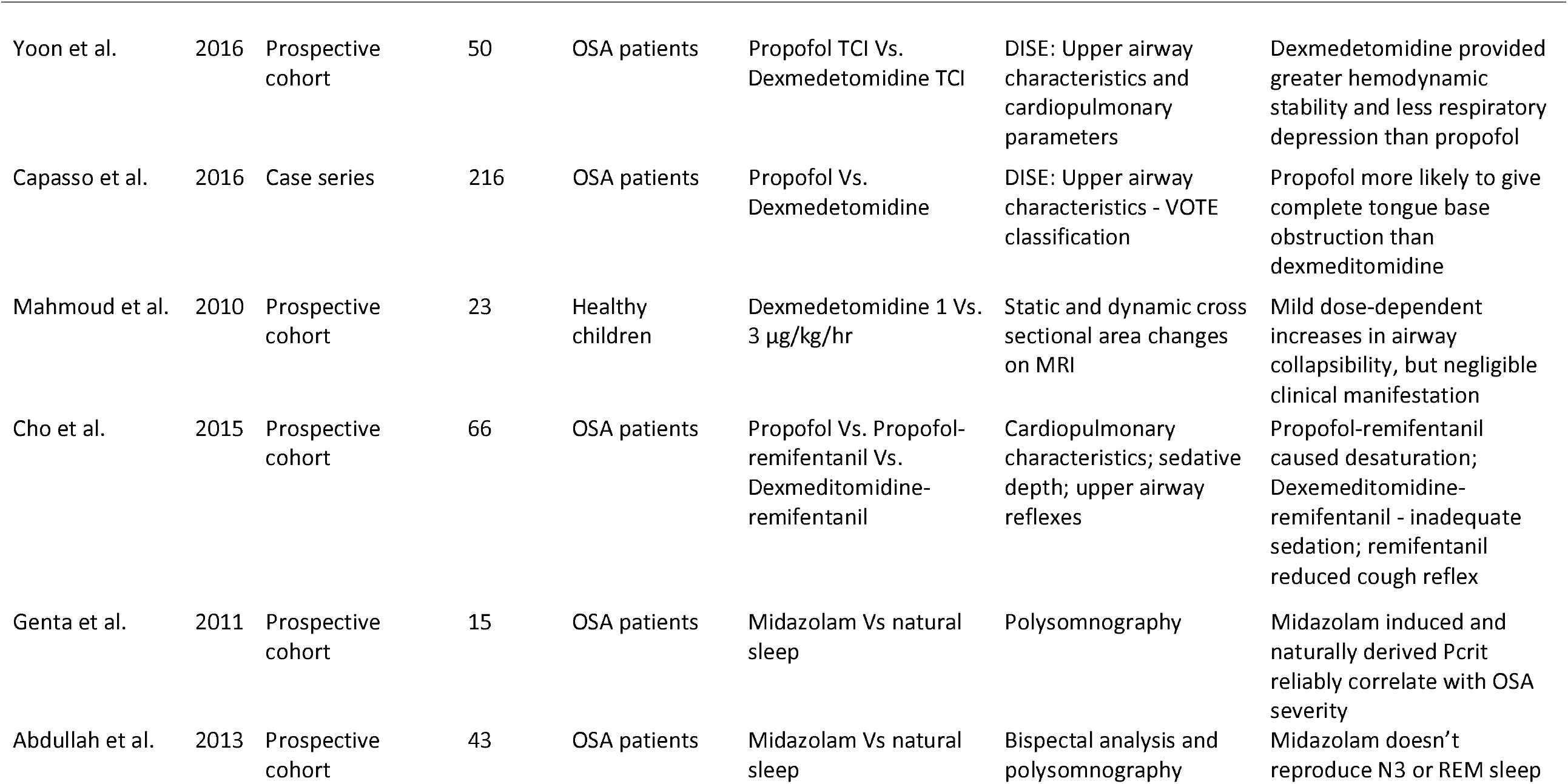
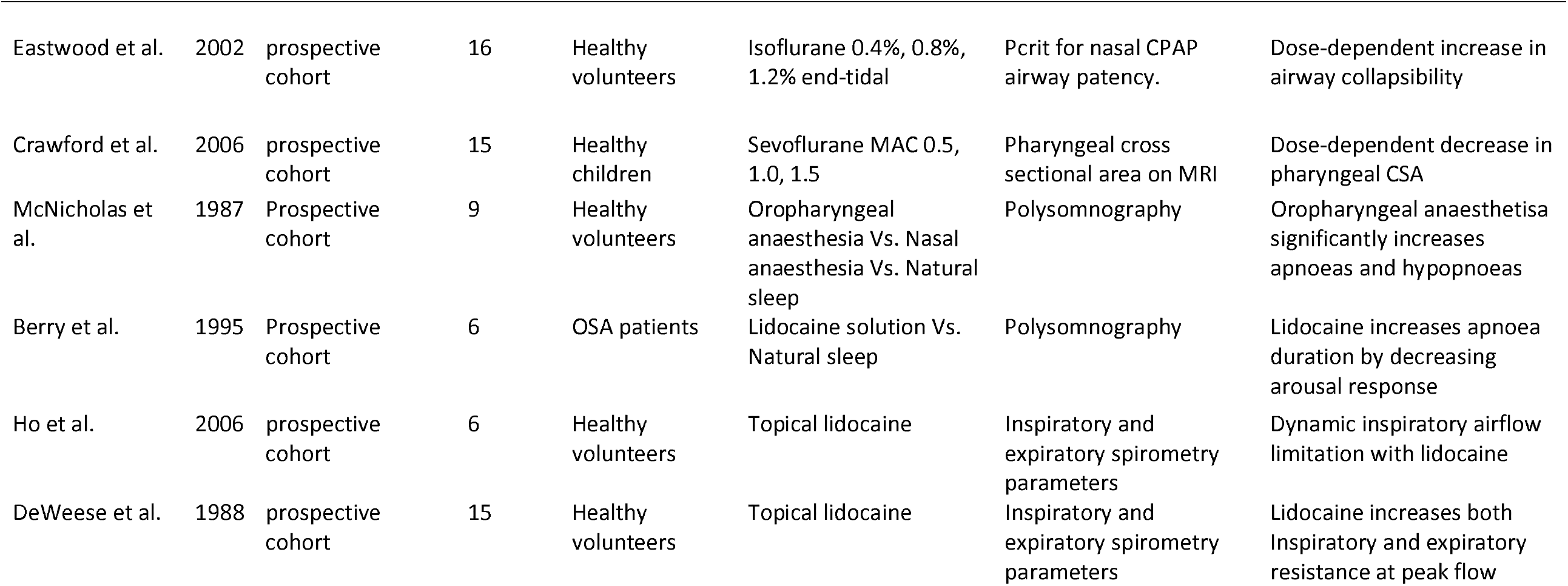
Summary of studies of the effect of anaesthetic agents on the upper airway reviewed

| Author | Year | Design | Number | Inclusion criteria | Intervention | Outcomes | Conclusion |
| --- | --- | --- | --- | --- | --- | --- | --- |
| Berry et al. | 2005 | Prospective cohort | 97 | OSA patients and healthy controls | Propofol | Induction of snoring or obstruction | Propofol did not induce snoring/obstruction in non-snoring controls but it was seen in OSA patients |
| Hillman et al | 2009 | Prospective cohort | 9 | Healthy volunteers | Step-wise induction with Propofol (0.5, 1.0, 1.5, 2.0, 2.5, 3.0, 4, 6.0 µg/ml TCI) | Genioglossus electromyography; Pcrit for CPAP airway patency | Dose dependent increase in airway collapsibility after transition from conscious to unconscious sedation |
| Rabelo et al. | 2010 | Prospective cohort | 15 | OSA patients and healthy controls | Propofol Vs. Natural sleep | Polysomnography and DISE | Main respiratory parameters are maintained with propofol but sleep architecture differs |
| Evans et al. | 2003 | Prospective cohort | 15 | Healthy children | Propofol 50 - 80 µg/kg/min | MRI: Cross sectional area at level of soft palate, tongue base, epiglottis | Dose-dependent decrease in CSA - worst at hypopharynx |
| De Vito et al. | 2011 | Prospective cohort | 40 | OSA patients | Propofol bolus Vs Propofol TCI | Cardiopulmonary characteristics; Upper airway characteristics - VOTE classification | Propofol TCI gave greater stability, safety and accuracy |
| Mahmoud et al. | 2013 | Prospective cohort | 60 | OSA patients | Propofol (100 vs. 300 µg/kg/m) Vs. Dexmedetomidine (1 vs. 3 µg/kg/hr) | cross sectional area on MRI | Non-significant dose dependent changes in both |
| Mahmoud et al. | 2009 | Retrospective descriptive | 82 | OSA patients | Propofol Vs. Dexmedetomidine | Successful completion of MRI, artificial airway required, additional airway manoeuvres required | Dexmedetomidine yielded more successful studies with less need for airway intervention |
| Yoon et al. | 2016 | Prospective cohort | 50 | OSA patients | Propofol TCI Vs. Dexmedetomidine TCI | DISE: Upper airway characteristics and cardiopulmonary parameters | Dexmedetomidine provided greater hemodynamic stability and less respiratory depression than propofol |
| Capasso et al. | 2016 | Case series | 216 | OSA patients | Propofol Vs. Dexmedetomidine | DISE: Upper airway characteristics - VOTE classification | Propofol more likely to give complete tongue base obstruction than dexmedetomidine |
| Mahmoud et al. | 2010 | Prospective cohort | 23 | Healthy children | Dexmedetomidine 1 Vs. 3 µg/kg/hr | Static and dynamic cross sectional area changes on MRI | Mild dose-dependent increases in airway collapsibility, but negligible clinical manifestation |
| Cho et al. | 2015 | Prospective cohort | 66 | OSA patients | Propofol Vs. Propofol-remifentanil Vs. Dexmedetomidine-remifentanil | Cardiopulmonary characteristics; sedative depth; upper airway reflexes | Propofol-remifentanil caused desaturation; Dexmedetomidine-remifentanil - inadequate sedation; remifentanil reduced cough reflex |
| Genta et al. | 2011 | Prospective cohort | 15 | OSA patients | Midazolam Vs natural sleep | Polysomnography | Midazolam induced and naturally derived Pcrit reliably correlate with OSA severity |
| Abdullah et al. | 2013 | Prospective cohort | 43 | OSA patients | Midazolam Vs natural sleep | Bispectral analysis and polysomnography | Midazolam doesn't reproduce N3 or REM sleep |
| Eastwood et al. | 2002 | prospective cohort | 16 | Healthy volunteers | Isoflurane 0.4%, 0.8%, 1.2% end-tidal | Pcrit for nasal CPAP airway patency. | Dose-dependent increase in airway collapsibility |
| Crawford et al. | 2006 | prospective cohort | 15 | Healthy children | Sevoflurane MAC 0.5, 1.0, 1.5 | Pharyngeal cross sectional area on MRI | Dose-dependent decrease in pharyngeal CSA |
| McNicholas et al. | 1987 | Prospective cohort | 9 | Healthy volunteers | Oropharyngeal anaesthesia Vs. Nasal anaesthesia Vs. Natural sleep | Polysomnography | Oropharyngeal anaesthesia significantly increases apnoeas and hypopnoeas |
| Berry et al. | 1995 | Prospective cohort | 6 | OSA patients | Lidocaine solution Vs. Natural sleep | Polysomnography | Lidocaine increases apnoea duration by decreasing arousal response |
| Ho et al. | 2006 | prospective cohort | 6 | Healthy volunteers | Topical lidocaine | Inspiratory and expiratory spirometry parameters | Dynamic inspiratory airflow limitation with lidocaine |
| DeWeese et al. | 1988 | prospective cohort | 15 | Healthy volunteers | Topical lidocaine | Inspiratory and expiratory spirometry parameters | Lidocaine increases both Inspiratory and expiratory resistance at peak flow |

Berry et al. 2005(11) found propofol to give data representative of OSA pathology and that it did not induce artificial obstruction in healthy volunteers. Of all the patients with OSA and snoring reported from collateral histories, desaturations and snoring was observed whereas this was not seen in any of the cases with healthy controls who were not previous snorers. Rabelo et al. (12) reproduced this finding but showed that propofol infused by TCI at 0.5 μg/mL higher than that needed to obtain procedural sedation exacerbated the magnitude desaturations seen in OSA patients compared with natural sleep, although maintains overall respiratory characteristics. TCI was also found to confer greater airway and cardiopulmonary stability compared with bolus dosing of propofol(13). Evans et al. (14) observed a dose dependent relationship with propofol and the extent of airway collapse in healthy children undergoing MRI. Cross sectional dimensions in the upper airway were reduced by propofol throughout as the propofol dose was increased, this was most profound at the level of the epiglottis in the hypopharynx and most marked following the transition from conscious to unconscious sedation(15).

Three papers from a group led by Mahmoud(16–18) used MRI to assess upper airways and commented on clinical correlation following escalating doses of dexmedetomidine (DEX) and propofol. In two instances the group found that whilst both DEX and propofol led to mild (statistically insignificant) changes in airway dimensions with escalating doses of each agent, there was little or no clinical effect in terms of cardiopulmonary parameters. In a third study they did however note that the need to perform airway interventions was more frequent with propofol than with DEX thus yielding more successful complete imaging sequences with the latter agent. These three studies found both DEX and propofol gave adequate sedation for non-invasive procedural anaesthesia needed for MRI. They did on several, though not statistically significant, occasions have to exclude patients from the DEX cohort as they required additional sedation with propofol for successful completion of the MRI. Yoon et al. (19) provided evidence that in patients undergoing DISE, whilst OSA findings correlated well between propofol and DEX, propofol gave greater haemodynamic instability and respiratory depression than DEX. In agreement with this, Capasso et al. (20)showed that propofol was more likely to induce complete airway obstruction at the level of the tongue base during DISE. Cho et al. (21) found that the addition of remifentanil to either DEX or propofol increased the severity of desaturations but did suppress the cough reflex which was assistive in completing the DISE study.

Two studies examined the effect of volatile anaesthetic agents on the upper airway. Crawford (22)demonstrates that reduction in upper airway cross sectional area is correlated at all anatomical levels with the depth of anaesthesia with sevoflurane. Studying subjects at MAC (minimum alveolar concentration) 0.5, 1.0 and 1.5 – however, none of these healthy children progressed to obstruction sufficient to cause desaturation. Eastwood’s group (23) showed that airway collapsibility also increased in a dose dependent way with isoflurane by measuring the inspiratory flows with a nasal continuous positive pressure system. The upstream pressure required to prevent upper airway obstruction was greater than atmospheric pressure.

Concerning the use of midazolam sedation, Genta et al. (24)found that it produced upper airway flow dynamics analogous to to those found during REM sleep in OSA patients. The P_crit_ (upstream airway pressure at which obstruction occurs) measured during midazolam sedation was equivalent to that obtained at the REM sleep stage – measured with polysomnography. However, Abdullah et al. (25) concluded using Bispectral analysis and polysomnography that midazolam sedation mimicked very little N3 sleep and no REM sleep – where most obstruction occurs but still support its use for accurately reflecting N1 and N2 sleep.

All four studies cited found that topical use of lidocaine in the asleep patient has deleterious effects on the airway including, increasing quantity and severity of desaturations through apnoea and hypopnoea(26–29). Also observed was an increase in airway resistance at both inspiratory and expiratory peak pressures. It is postulated that local anaesthetics reduce the upper airway reflexes which contribute to airway patency when sleeping.

### The Natural Sleep Induction Technique with Propofol

This article has reviewed anaesthetic agents given with the intention of provided drug induced sedation throughout the entirety of the procedure. A technique adopted by the senior author utilises propofol’s rapid onset and offset (30) clinical effect by first inducing a relatively deep sedation by providing a bolus of propofol. A flexible nasendoscope is then inserted into the correct position to examine the upper airway. Because of the deep sedation at this point, minimal sneezing or stimulation is apparent. No further propofol is given for the rest of the procedure. It takes approximately 2 to 5 minutes for the patient to reach a lighter level of sedation that is similar to that seen during TCI propofol techniques. The environmental conditions are altered in the theatre to ensure as little light and noise is generated around the patient. As the effects of propofol eventually become relatively subclinical, the patient is easily able to be woken up by gently calling out their name or something similar. Most patients however carry on sleeping naturally if the quiet environmental conditions are maintained. This easily rousable state is almost indistinguishable from natural sleep and the authors believe this accurately represents the conditions seen during naturally induced sleep.

The experience of the senior author gained from the first 21 patients with this new technique revealed a number of important advantages:

1. The entire range of sedation depths are evaluated giving a more complete representation of a patient’s natural sleep. Admittedly a large percentage of this is during a state under the direct influence of an anaesthetic agent. The senior author’s opinion is that there is probably a variation in airway collapsibility during a night of natural sleep, and therefore knowledge of this spectrum is important in the investigation of these OSA patients. There is the initial minute or so when the patient is too sedated and almost complete airway collapse is apparent in all patients. Appreciating that this is an artifactual collapse caused by the anaesthetic agent is key to not being deceived into making an inappropriate interpretation of the DISE. Typically, video recording of the investigation is initiated when the patient is making normal tidal volume breaths, which requires a certain amount of propofol washing out of the physiological system. This is continued until the patient is clearly naturally sleeping and can be woken up with minimal stimulation.
2. In this cohort there was no obvious difference in obstructive level seen during the commonly used sedation level obtained with TCI propofol and when the patient is assumed to be naturally sleeping at the end of the procedure. This we believe not only validates the use of Propofol in DISE, but also validates the use of DISE as an investigation that closely represents natural sleep.

The disadvantages of this technique are firstly that it certainly takes longer to perform than a normal DISE investigation. On average the whole procedure takes between 10 and 15 minutes using this technique. The authors believe that validating what is seen during the anaesthetically driven state provides invaluable information and reassurance that a true representation of natural sleep was captured. Secondly it appears to be harder for our anaesthetic colleagues to master when compared to TCI of propofol. It can be daunting for inexperienced practitioners to allow a patient to have an unsecured airway for a significant length of time. Thirdly this technique requires a certain amount of interpretation skill by the surgeon who is evaluating a number of different levels of sedation during one procedure. On two occasions patients woke up before an adequate examination of the airway was possible. In these situations, the process was repeated and good conditions were eventually achieved. Finally, if the bolus provided is too great, an overly deep sedation may occur making the procedure much longer as the patient would require manual mask ventilation and oxygen delivery whilst the anaesthetic effects dissipate. With experienced anaesthetic doctors, this however was not a feature in our cohort.

## Discussion

This review compares findings from studies examining the effects of a variety of sedating agents on the upper airway and also on cardiopulmonary parameters, both of which contribute significantly to successful completion of DISE leading to efficacious surgical planning. The aim of the use of the selected agent is to mimic natural sleep as closely as possible and in doing so reveal the sites and extent of obstruction occurring on a nightly basis in OSA patients(31). If the collapsibility produced by the agent is too exaggerated, the surgical intervention planned may be too extensive(32,33). Worse yet would be an agent which is selective in its site of obstruction in doing so giving a false positive on DISE and possibly leading to the wrong surgical procedure(34).

## Conclusion

This review concludes that whilst studies into the effect of anaesthetic agents on the upper airway and other parameters necessary for consideration in the successful completion of DISE are limited, there is good evidence for the efficacy for safe sedation using certain agents in presence of clinicians confident in airway management. There is no industry standard (35) or clear clinical guidance for the use of anaesthetic agent in DISE (36). All agents give elements of upper airway collapse and cardiopulmonary instability in varying degrees. Of the agents examined dexmedetomidine and propofol have been found to be relatively safe and reliable for conducting DISE. Most agents have been shown to have a dose dependent effect on airway collapsibility, necessitating careful titration of doses and attentive monitoring when considering when to obtain the diagnostic images needed for surgery aimed at correcting sleep disordered breathing.

The induction of natural sleep using propofol and the further validation of DISE is encouraging information. The authors aim to further evaluate this state after the clinical effects of propofol have dissipated with more objective testing.

## Data Availability

All data produced in the present work are contained in the manuscript

## Compliance with Ethical Standards

- Disclosure of potential conflicts of interest
  - Dr Oliver Sanders declares that he has no conflict of interest
  - Professor Bhik Kotecha declares that he has no conflict of interest
  - Mr Vik Veer declares that he has no conflict of interest
  - All authors declare they have no conflicts of interest
- Research involving human participants and/or animals
  - This article does not contain any studies with human participants or animals performed by any of the authors.
- Informed consent
  - Only studies where informed consent was obtained from participants were reviewed in this article

